# Impact of reproduction number on multiwave spreading dynamics of COVID-19 with temporary immunity : a mathematical model

**DOI:** 10.1101/2020.09.30.20204636

**Authors:** B Shayak, Mohit M Sharma, Manas Gaur, Anand Kumar Mishra

## Abstract

**Objectives:** The recent discoveries of phylogenetically confirmed COVID-19 reinfection cases worldwide, together with studies suggesting that antibody titres decrease over time, raise the question of what course the epidemic trajectories may take if immunity were really to be temporary in a significant fraction of the population. The objective of this study is to answer this question.

**Methods:** We construct a ground-up delay differential equation model tailored to incorporate different kinds of immune response. We consider two immune responses here : (*a*) short-lived immunity of all kinds, and (*b*) short-lived sterilizing immunity with durable severity-reducing immunity.

**Results:** Multiple wave solutions to the model are manifest for intermediate values of the reproduction number *R*; interestingly, for sufficiently low as well as sufficiently high *R*, we find conventional single-wave solutions despite the temporary immunity.

**Conclusions:** The versatility of our model and its very modest demands on computational resources ensure that a set of disease trajectories can be computed virtually on the same day that a new and relevant immune response study is released. Our work can also be used to analyse the disease dynamics after a vaccine is certified for use and information regarding its immune response becomes available.

**Highlights:**

- We show a subtle interplay between public health measures and immune response.
- We account for different durations of sterilizing and severity-reducing immunity.
- Our results prepare us for what might happen with large-scale temporary immunity.
- Our model is relevant for predicting disease dynamics after a vaccine is invented.

## Introduction

Between 24^th^ August and today, nine phylogenetically confirmed cases of COVID-19 reinfection have been discovered worldwide – the first in Hong Kong (To et. al. 2020), two more in Greater Noida, India (Sinha, Gupta et. al. 2020), two in Belgium (van Elslande et. al., Selhorst et. al. 2020), two in USA (Goldman et. al., Tillett et. al. 2020), one in Ecuador (CGTN 2020) and one in the Netherlands (Mulder et. al. 2020). In eight of these cases, the patients were presumed immunocompetent; among these, the second infections were milder than the first in six cases and more severe in two. Much about the immune response to SARS-CoV-2 is currently unknown. Edridge et. al. (2020) have found that immunity against infection by benign coronaviruses (not SARS-CoV, MERS-CoV or SARS-CoV-2!) lasts for a few months and reinfection is common from one year onwards. Wajnberg et. al (2020), at Mount Sinai Hospital in New York, USA found that among a cohort of almost 20,000 patients, all but one demonstrated significant antibody titre levels in blood plasma at three months following the original infection. Siddiqui et. al. (2020) corroborated these results via a smaller study conducted at Max Hospital, New Delhi, India.

An observational cohort study (Crawford et. al. 2020) on 34 patients conducted at the University of Washington at Seattle found that over a 3- to 5-month period, antibody concentrations decreased over time, in a manner consistent with the immune responses to acute infections by other viruses including influenza, SARS and MERS. In these infections, the initial decrease in titres is followed by a plateau; whether this is true of SARS-CoV-2 is unknown as of now. Abu-Raddad et. al. (2020) have reported that out of more than 1,30,000 patients in Doha, Qatar infected with COVID-19, 243 persons reported a positive swab 45 or more days after the original positive test; 54 out of these 243 had “strong or good evidence for reinfection”. All the reinfections were asymptomatic or presented mild symptoms on the second bout; however the initial symptom profile of these patients had been mild or asymptomatic as well. Kowitdamrong et. al. (2020) report upto 20 percent post-infection seroconversion failure rate in mild or asymptomatic patients in Thailand – they do not document any reinfection cases. Lumley et. al. (2020) have very recently reported an antibody half-life of 85 days and a median time of 137 days to loss of positivity; these results are corrborated by Robertson et. al. (2020). A few potential cases of reinfection have also been reported in media for some time before the first documented case – in these instances, the evidence is not fully credible (McCamon, Saplakoglu, Ackerly 2020).

Two systematic reviews deal with the antibody response (Post et. al. 2020) and the cellular immune response (Shrotri et. al. 2020) to the virus. The former contains amplification of the kind of results we discussed above; the latter states that the response of T-cells to SARS-CoV-2 is currently unknown. Lavine et. al. (2020) propose that the immune response to the virus will determine the manner of transition of COVID-19 to endemicity; in the absence of known facts, this work is predominantly speculative at this time. Recent cross-sectional studies from Paris (Anna et. al. 2020) and London (Ward et. al. 2020) find decreasing seroprevalence as a function of time; this may be related to decreasing titre levels and might indicate a possibility of reinfection. In two recent, reassuring developments, Zuo et. al. (2020) in Manchester, UK and Ogega et. al. (2020) in Baltimore, USA have found significant levels of T-respectively B-cells in recovered patients at 25 respectively 15 weeks following infection; both studies included mild or asymptomatic patients as well as patients with very low antibody titre levels. This indicates that durable cellular immunity may be present against this virus even if antibodies decay over time.

Although the reinfection cases so far are isolated, the almost daily updates on the immune response to SARS-CoV-2 make us wonder what the epidemiological consequences might be if the immunity duration indeed turns out to be finite for a significant fraction of the population. The only approach which can allow inroads into this question is mathematical modeling. Giordano et. al. (2020) and Bjornstad et. al. (2020) account for the possibility of reinfection, with the latter finding an oscillatory approach towards the eventual endemic equilibrium. Kosinski (2020) however finds multiple waves of COVID-19 if the immunity duration is finite. Sandmann et. al. (2020), in an analysis of this situation in the context of a vaccine, find smooth oscillations about an endemic equilibrium, which change to jerky oscillations with periodic lockdown. In this Article, we show that the case trajectories with temporary immunity actually depend in an intricate manner on the reproduction number *R*. Before commencing our analysis, we clarify that we are currently treating large-scale temporary immunity as a hypothetical, hopefully worst-case scenario, regarding the validity of which we do not yet have sufficient available data. Also, in this work, we do not attempt to model the effects of vaccination; all the possibilities that might come into play after introduction of this new variable will be considered in a subsequent analysis.

## Methods

We start from a delay differential equation (DDE) model constructed by our group (Shayak et. al. 2020, Mohit and Shayak 2020, hereinafter referred to as “References [26,27]”), which accounts for many realistic features associated with COVID-19 transmission. This model can be easily adapted to accommodate a given immune response. The dependent variable *y*(*t*) denotes the cumulative number of corona cases in the region of interest (typically a town, neighbourhood, village or other area with good interaction among the inhabitants) as a function of time, measured in days. The parameters in the model are:

- *m*_0_ : the per-case spreading rate which accounts for factors such as the degree of mobility (i.e. lockdown/unlock), extent of mask use, extent of handwashing and other public health measures
- *τ*_1_ : the asymptomatic transmission period which we take to be 7 days throughout
- *τ*_2_ : the pre-symptomatic transmission period which we take to be 3 days throughout
- *μ*_1_ : the fraction of patients who are asymptomatic (0<*μ*_1_<1)
- *μ*_3_ : the fraction of patients (both symptomatic and asymptomatic) who are NOT detected in contact tracing drives and quarantined (0<*μ*_3_<1)
- *N* : the total number of susceptible people in the region at the start of the epidemic

In References [26,27], we had assumed that one bout of infection renders everlasting immunity (practically, immunity lasting longer than the epidemic’s progression). Now, we account for two different kinds of immune response, as follows.

- Simple response: In this scenario, one bout of infection renders a person insusceptible to fresh infection for a time *τ*_0_ days (*τ*_0_ much greater than *τ*_1_ and *τ*_2_) following recovery, after which s/he again turns susceptible to the disease in its original form.
- Complex response: In this scenario, adapted from Lavine et. al. (2020), a person is initially susceptible to the current, highly virulent form of the disease. The first bout of infection renders him/her completely insusceptible for *τ*_0_ days following recovery, after which s/he turns susceptible to a lower virulence form of the disease. If infected the second time, then s/he becomes permanently insusceptible to further infection.

As is customary in lumped parameter or compartmental models, the value of *τ*_0_ used in the model must be an average over the entire population. The experiences of the first few confirmed reinfections may well indicate a general trend favouring the second immune response to the first; in the absence of data, we shall work with both assumptions.

It can be shown (Section 2 of the Supplementary Material) that with the simple immune response, the dynamics of the disease is governed by the DDE

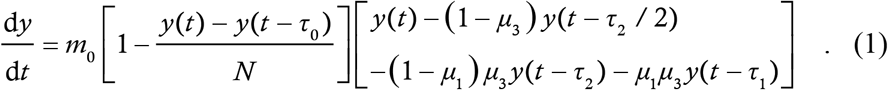

With the complex immune response, we need two dependent variables : *y*(*t*), the cumulative number of cases of the high virulence form and *z*(*t*), the cumulative number of cases of the lower virulence form. We let *μ*_1*a*_ and *μ*_1*b*_ (presumably greater than *μ*_1*a*_) be the asymptomatic fractions for the two forms and keep all other parameters identical for both. Then, the dynamics of *y* and *z* are governed by the following coupled system of DDEs

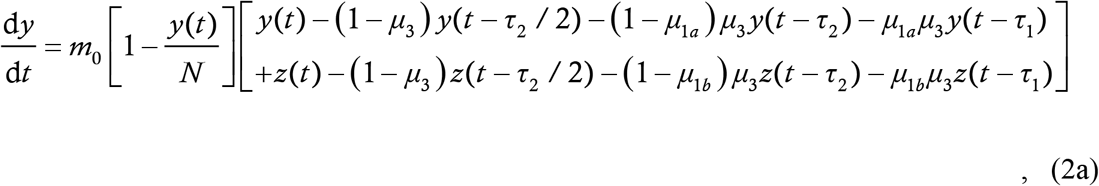

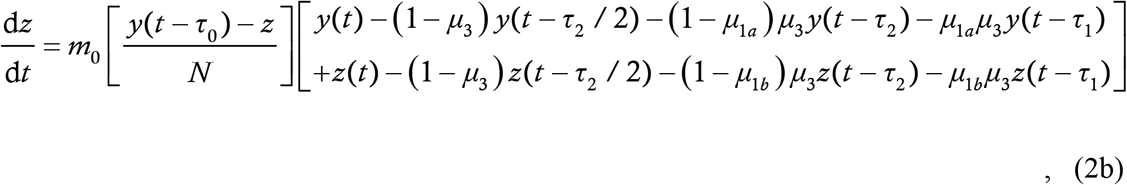

whose derivation is again given in the Supplementary Material, Section 2.

For both models, we present the solutions in a Notional City having an initial susceptible population of *N*=3,00,000 (the epidemic durations and case trajectory shapes are almost independent of *N* and the case counts scale as *N* for large *N*) and 80 percent asymptomatic carriers i.e. *μ*_1_=0·8 (or *μ*_1*a*_=0·8 if appropriate) in the current, high virulence form of the disease. We let *μ*_1*b*_=0·95 for the lower virulence form and take the immunity duration *τ*_0_ to be 200 days. We solve the equations numerically in Matlab using 2^nd^ order Runge Kutta method with step size 0·001 day. The initial condition function we take is zero cases for the first 193 days followed by linear growth of cases at 100 cases/day for the next seven days (with the complex immune response, this growth refers to the high virulence cases – the lower virulence cases remain zero). We take *t*=0 to be the 194^th^ day of the initial condition period.

One issue needs special mention – in a numerical simulation, the case rate will never be identically zero but will be something like 0·001 cases/day (or machine epsilon cases/day). This can pose a serious problem in a situation where there are potential second waves of the epidemic. To circumvent this issue, we have arranged for manual termination of the run if the case rate becomes sufficiently low. Defining the number of active cases at time *t* to be *y*(*t*)−*y*(*t*−14), we stop the run if there is less than one active case for 14 consecutive days. While the number 14 (twice) may be somewhat arbitrary, the criterion of a low enough active case count for a long enough period is a very reasonable indicator of the true end of the outbreak. We run all simulations either upto *t*=1400 days or until they terminate, whichever is earlier.

## Results

All results in this Section are based on an assumed immunity period of 200 days. The exact value is not of the greatest significance; as we shall see, the important thing is whether this period is shorter or longer than the evolution of the outbreak with permanent immunity. Six different solution classes corresponding to different parameter values – labelled as Notional Cities A through F – of the model with permanent immunity have been derived in References [26,27] and recapitulated in Section 1 of the Supplementary Material. We shall consider the same Cities in the present analysis.

### Scenario 1 : Simple immune response

We first consider the simple immune response, modelled by (1). Notional City A has *m*_0_=0·23 and *μ*_3_=0.5, which, with permanent immunity, resulted in the epidemic’s being driven monotonically to containment in 120 days. With the simple immune response, the time-trace of the disease is shown in Figure 1 below. Here and henceforth, we show three things in the same plot : *y*(*t*) as a blue line, its derivative as a green line and the “epi-curve” or weekly increments in cases scaled down by a factor of 7, in grey bars.

**Figure 1:**
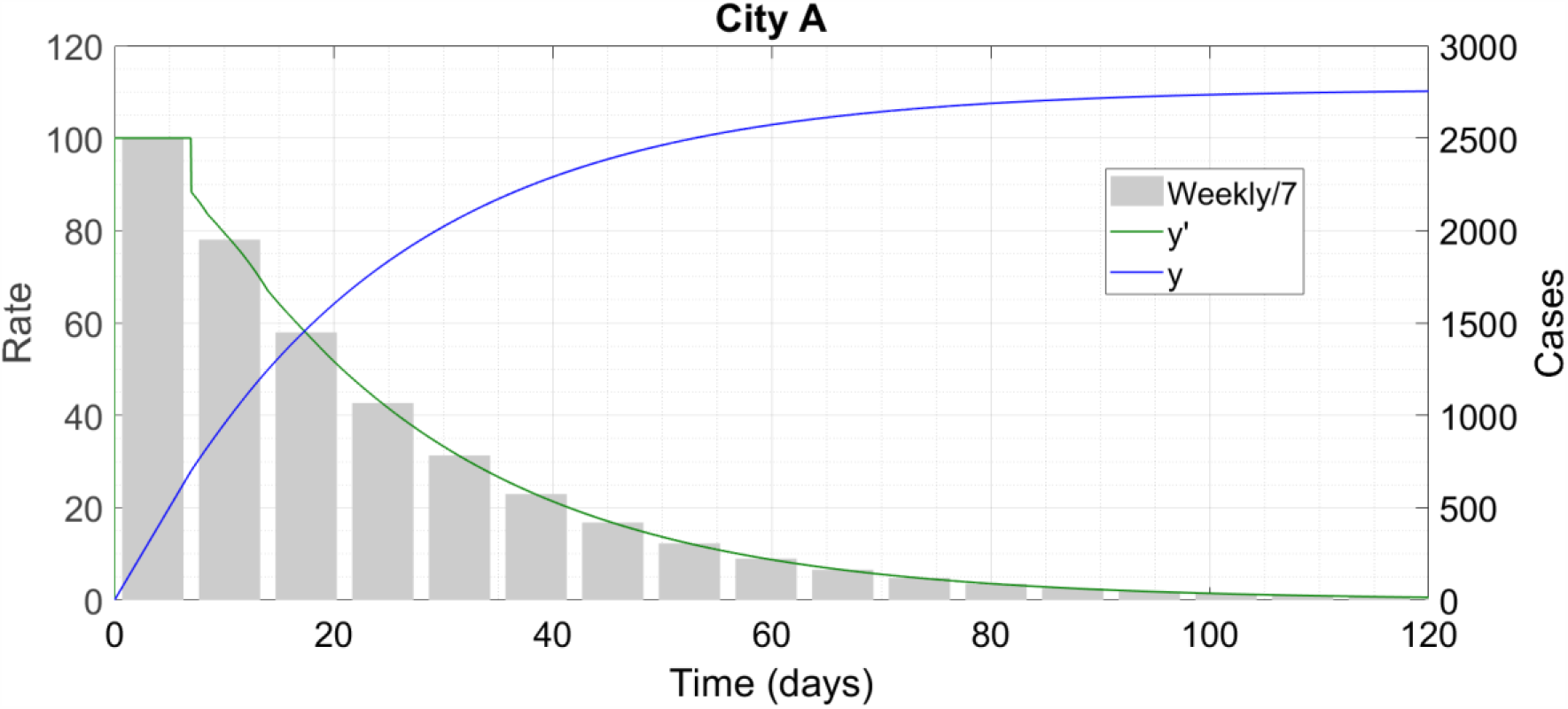
City A eliminates the epidemic via non-pharmaceutical interventions alone.

The response remains the same as it was with permanent immunity. Next, we present City C, which has *μ*_3_=3/4 and *m*_0_=0·5 all the time. (We use 0·23 for a “low” value of *m*_0_ since we obtained it from data fits in References [26,27]; the chosen “high” value of *m*_0_=0·5 generates 90 percent infection level at the end of the outbreak.) With the simple immune response, the trajectory is shown in Figure 2 below.

**Figure 2:**
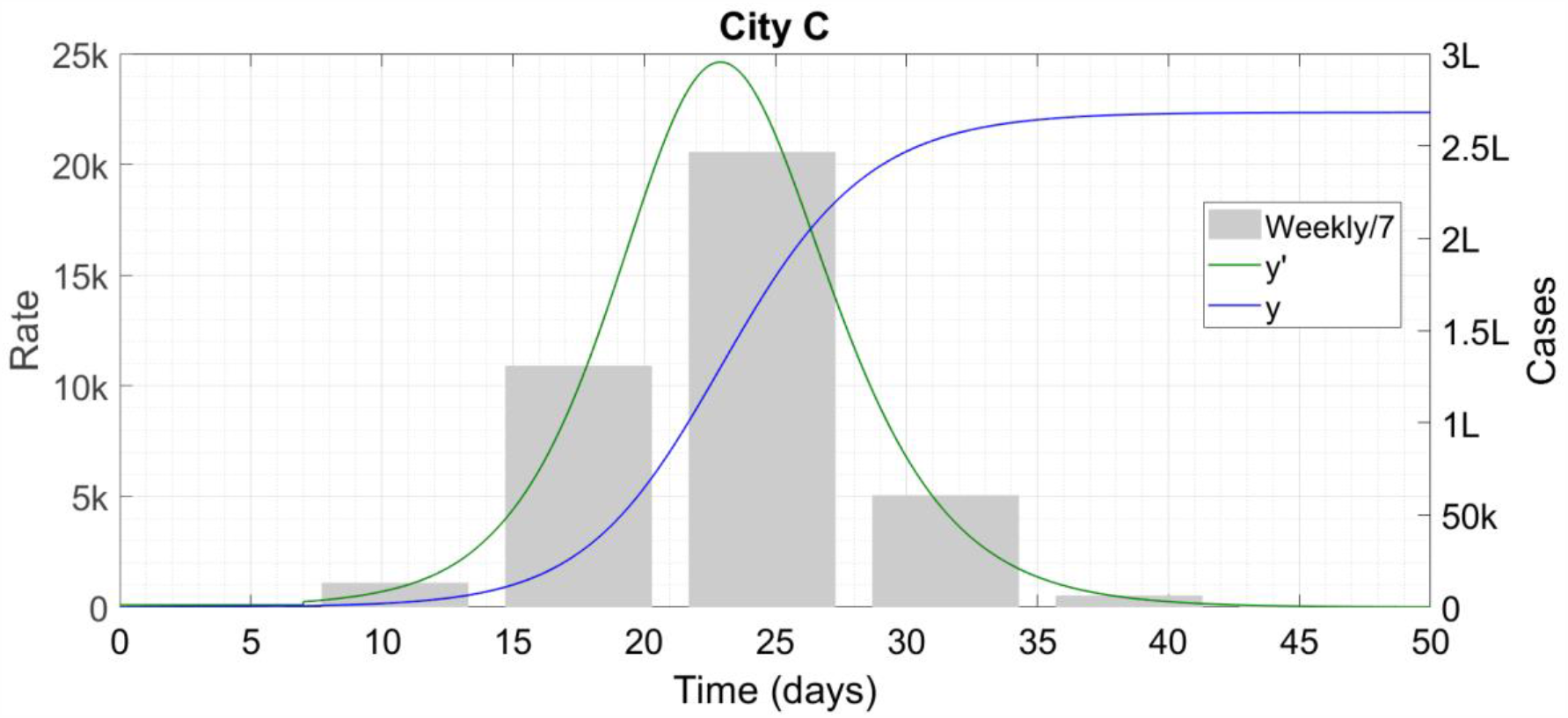
City C reaches herd immunity well before the 200-day mark. ‘k’ denotes thousand and ‘L’ hundred thousand.

Once again, there is no change in behaviour from the permanent immunity case. City B has *m*_0_=0·23 like City A but *μ*_3_=3/4 like City C. With permanent immunity, City B crawls up to about 26 percent infection level over a period of 220 days. Here however, the 220-day run becomes longer than the assumed immunity duration of 200 days, and the result, now labelled as City B1, is shown in Figure 3.

**Figure 3:**
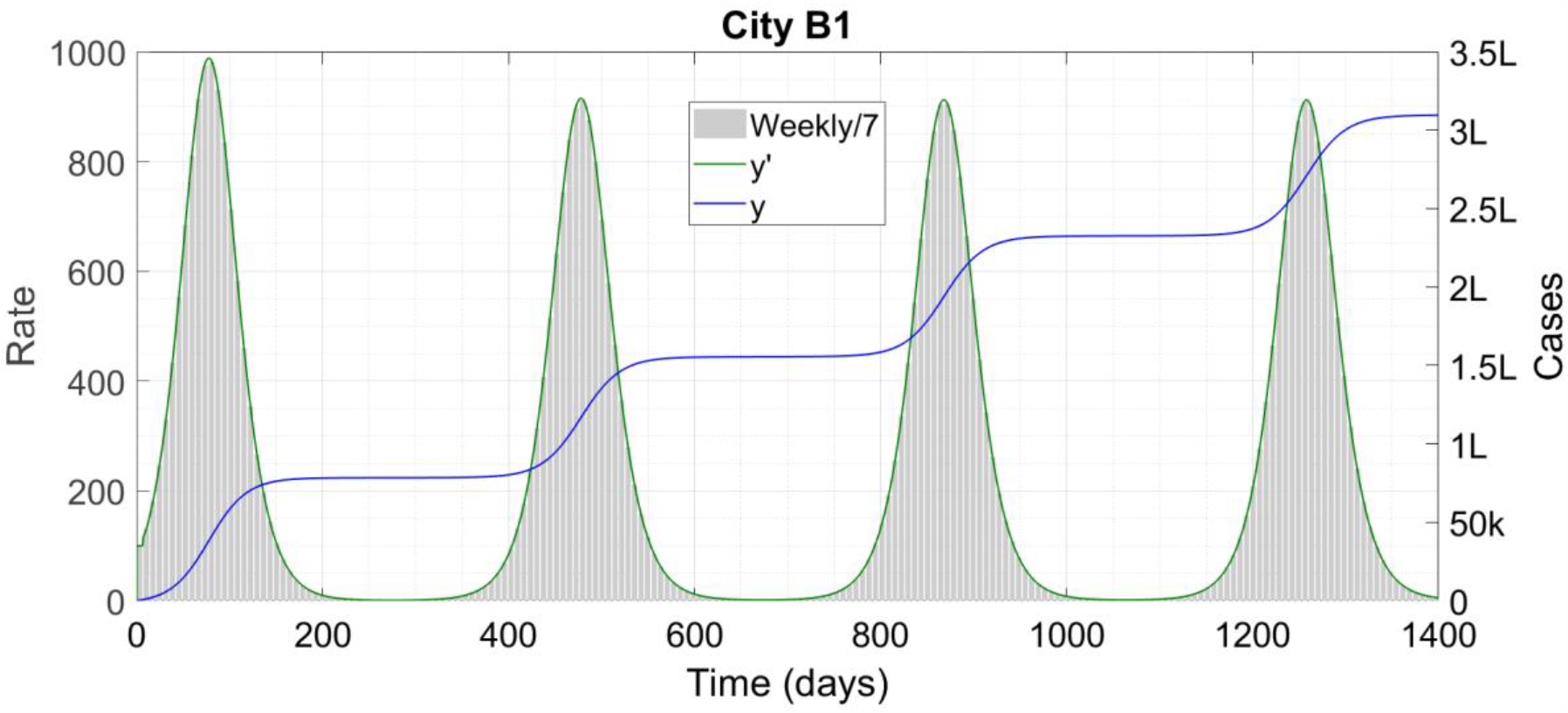
City B1 (we shall use B2 to denote the same city with complex immune response) is a less effective form of A and has multiple waves of COVID-19. ‘k’ denotes thousand and ‘L’ hundred thousand. We have stopped the simulation at 1400 days – the waves persist after that time as well.

We can see wave after wave of outbreaks here. While we do not believe that the disease can really run unmitigated for four years, the long simulation runtime shows the perfect periodicity of the case trajectories. The case count at the end of the run is greater than the city’s initial susceptible population, so at least some people have been infected at least twice. Cities D and E of References [26,27] were similar to C and A respectively and again, they do not show any change when the new immune response is incorporated. City F, which demonstrates reopening-induced second waves, is more interesting. We start off this City with the parameter values of B, on a path to multiple waves. One hundred and fifty days into the outbreak it reopens completely, raising *m*_0_ to 0·5, aiming to infect its entire population before immunity runs out. The result is shown in Figure 4.

**Figure 4:**
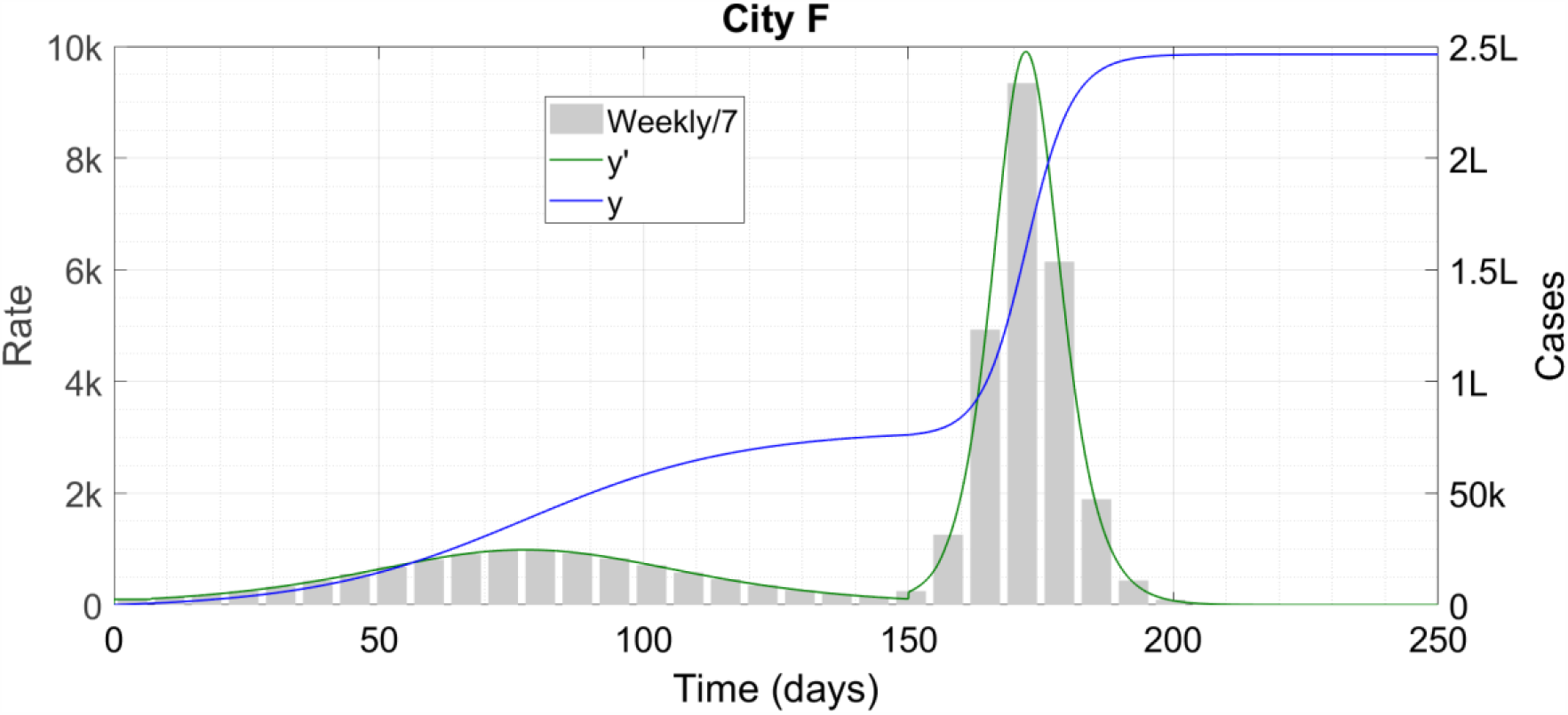
City F is bold and “beats the virus to the finish line”. ‘k’ denotes thousand and ‘L’ hundred thousand.

Howsoever controversial this strategy might appear, it works. The reopening generates an immediate second wave but it does succeed in making the epidemic vanish completely at 210 days and 2,46,000 infections. This strategy however has an unhappy variant. City G, instead of reopening at one shot, increases *m*_0_ linearly from 0·23 to 0·50 over the interval from 150 to 250 days. The resulting infection profile is shown in Figure 5.

**Figure 5:**
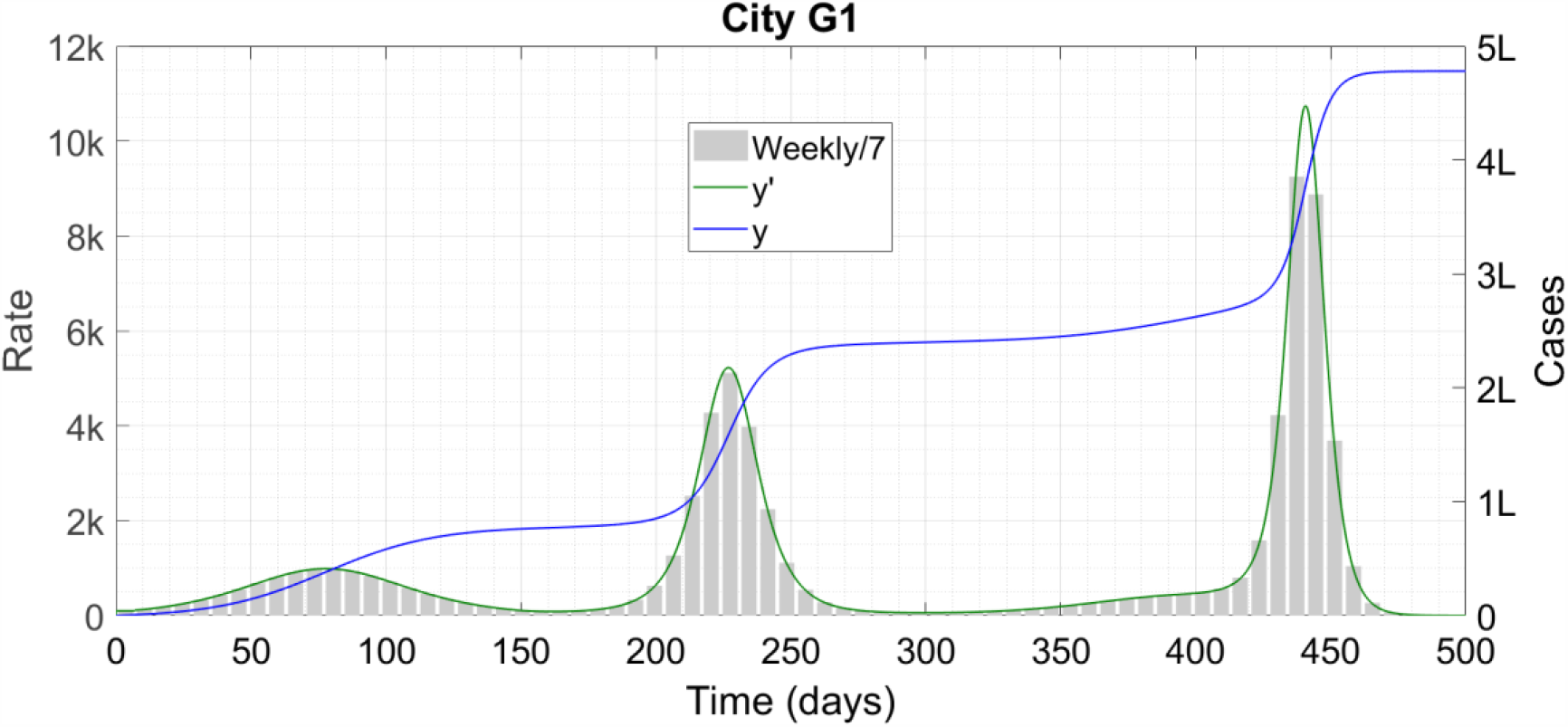
City G1 is F gone wrong, and the error carries a price. ‘k’ denotes thousand and ‘L’ hundred thousand.

There are three separate waves of infections here, each more severe than the first, and the cumulative case count exceeds 150 percent of the total population.

### Scenario 2 : Complex immune response

We now consider the complex immune response, modelled by (2). Numerical work confirms our expectation that Cities A, C and F will show no difference from the simple immune response case since the epidemic terminates before the immunity duration lapses. We focus on City B, now renamed to B2. For this plot, the colour legend will be *y* in blue, *y* in green, *z* in red, *z* in magenta and the epi-curves in grey for *y* and cyan for *z*. The results are in Figure 6.

**Figure 6:**
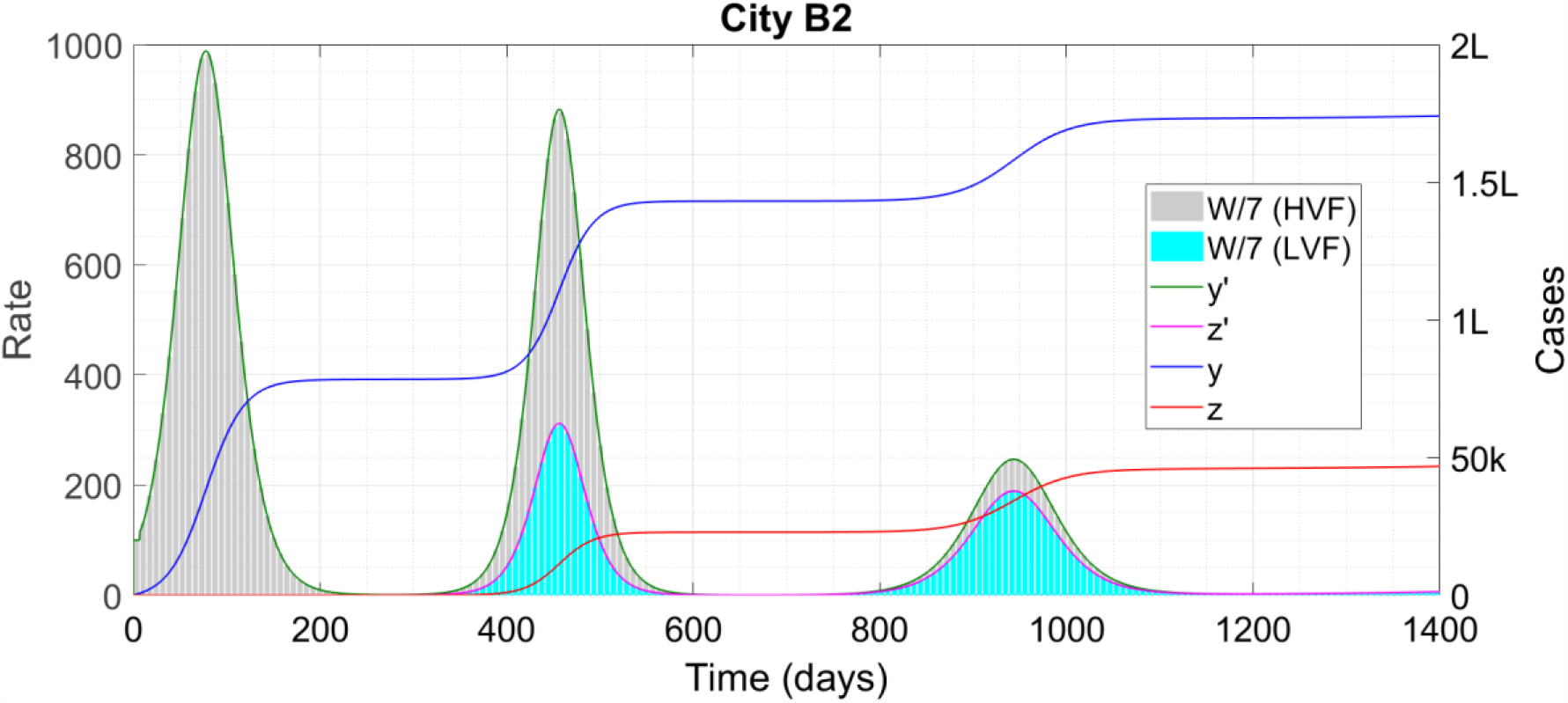
City B with complex immune response. ‘k’ denotes thousand and ‘L’ hundred thousand. W/7 denotes the weekly increments in cases scaled down by 7 as previously, while HVF and LVF refer to the high and lower virulence forms of the disease respectively.

Again there are multiple waves, but this time they are progressively attenuated so that the total fraction of the high virulence infection remains less than 60 percent. An equally significant difference between the two immune responses occurs with City G, Figure 7 below.

**Figure 7:**
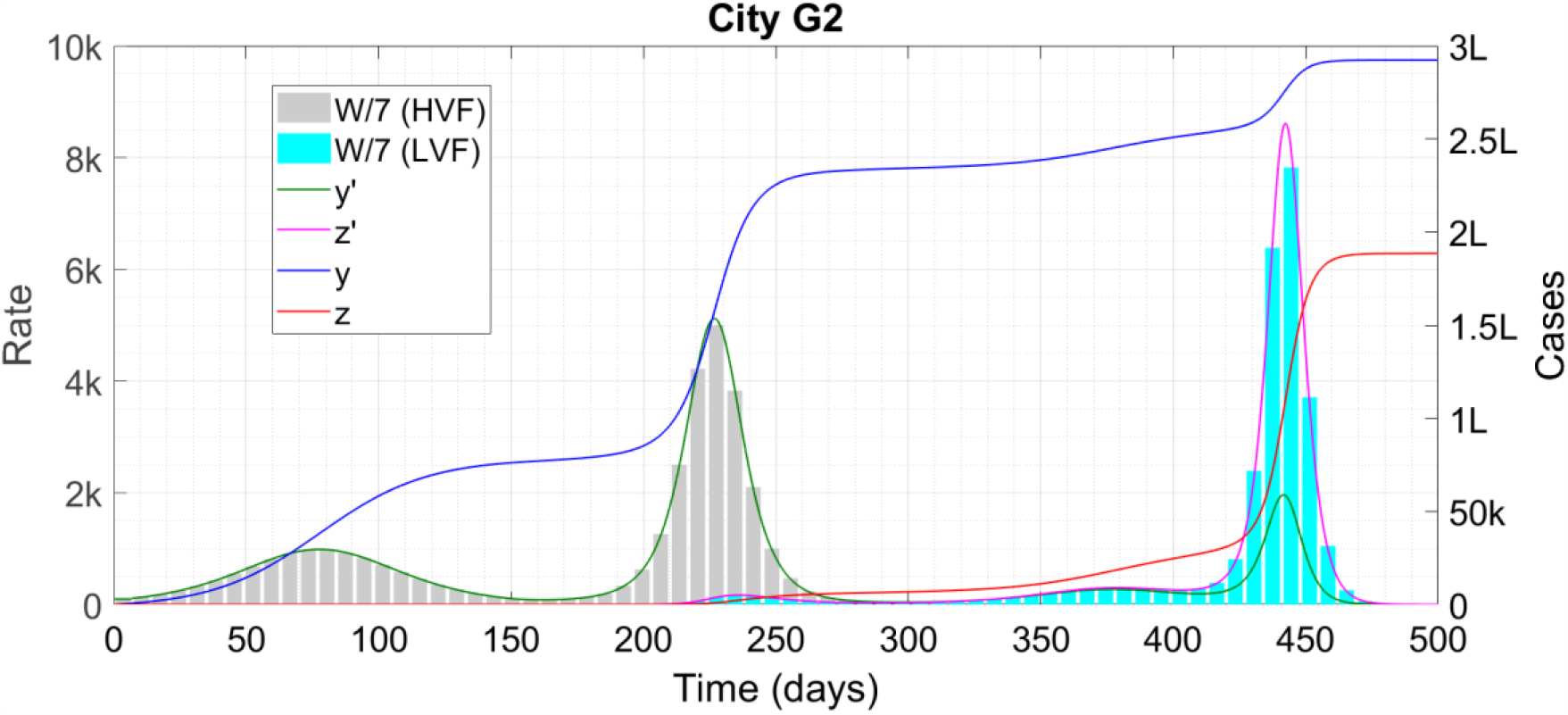
City G with complex immune response. In the right third of the plot, the cyan bars obscure the smaller grey bars; the latter are enveloped by the green curve which is still visible. ‘k’ denotes thousand and ‘L’ hundred thousand. W/7 denotes the weekly increments in cases scaled down by 7 as previously, while HVF and LVF refer to the high and lower virulence forms of the disease respectively.

The massive third wave of G1 now occurs almost entirely in the lower virulence form.

## Discussion

The difference between the cities A, B and C lies in their reproduction number *R*, which is proportional to *m*_0_ if other parameters are held constant. City A has the starting value *R*_0_=0·886, City B has *R*_0_=1·16, and City C has *R*_0_=2·5. With permanent immunity, a low but greater-than-unity *R*_0_ leads to a long epidemic duration with a lower caseload, while high *R*_0_ leads to a shorter duration with a higher caseload. The latter carries a significant risk of overwhelming healthcare facilities and causing unnecessary deaths. With temporary immunity however, we see multiple waves of disease if the cutoff interval *τ*_0_ becomes less than the evolution time with permanent immunity (Supplement, Section 3 contains some pedagogical examples that motivate these findings).

In this example, we took *τ*_0_ to be 200 days, which is towards the longer end of the free evolution duration with time-independent parameters (with time-dependent parameters like periodic lockdown, the epidemic durations can become much longer). Hence we see the wave solutions in a small region of parameter space – we find that with the simple immune response, for all parameters except *m*_0_ being held to their City B values, a containment (City A) solution for low *m*_0_ gives way to a multiple wave (City B1) solution as *m*_0_ is increased above 0·20, while that cedes to a one-shot logistic-like (City C) solution at *m*_0_=0·25 and higher. A shorter immunity duration would have caused multiple waves over a larger range of *m*_0_. In our example, the waves are close to sinusoidal near *m*_0_=0·20; as *m*_0_ is increased, the crests get higher and narrower and the troughs lower and wider. From a public health perspective, a low and wide trough might be an opportunity to intensify test-trace-treat efforts and stamp the disease out. With the complex immune response, the total numbers of cases are bounded by 3,00,000 infections of both *y* and *z*. Hence, the infinitely periodic waves are no longer possible – they keep attenuating in size before the epidemic terminates outright. In both Cities B and G, the time interval between successive waves appears to be in the range 1·5*τ*_0_ to 2*τ*_0_. Cities F and G show the possible scenarios with reopening. In Table 1, we summarize the different case trajectories possible for different values of *R* with three kinds of immune response – permanent, simple and complex.

**Table 1:**
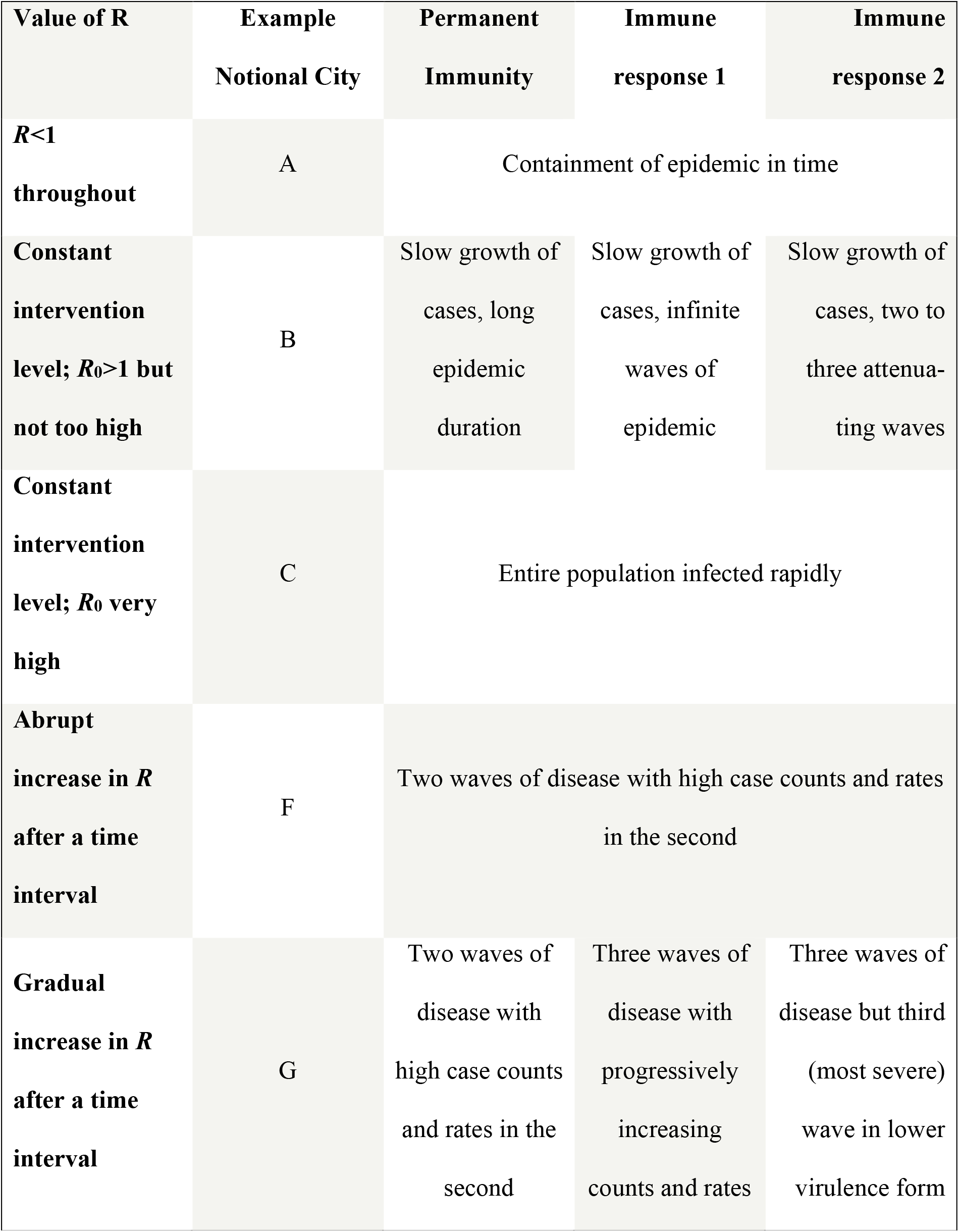
Different scenarios possible with different levels of intervention combined with different kinds of immune response.

We note that the second waves seen after relaxing public health interventions are fundamentally different from the second and subsequent waves exhibited under the aegis of temporary immunity. The former is caused by more susceptible people getting exposed and infected, as in the second wave of City F, while the latter is caused by recovered cases turning re-susceptible and thus increasing the size of the susceptible pool, like the waves in City B1. The waves in City G1 are of both types, with the second wave being of the former type and the third of the latter. This is why in City G2 (Figure 7), where previously infected people suffer a “different” disease, the second wave occurs primarily in the high virulence form while the third occurs in the lower virulence form. The phenomenon we are currently seeing in European countries and elsewhere is overwhelmingly the result of weakening of public health intervention measures and not of immunity having run out. We are unsure as to what factors governed the successive waves of the deadly 1918-9 influenza pandemic and how that disease eventually ended.

Briefly revisiting the prior Literature, we find that some prior works (Giordano et. al., Bjornstad et. al. 2020, Cooke et. al. 1996, Travicki 2017, Kiran et. al. 2020) appear to have missed the multiple wave solutions possible with temporary immunity. While Kosinski (2020) and Sandmann et. al. (2020) do find these waves, they do not obtain the diverse possible epidemic trajectories depending on *R*. Since *R* is governed by public health interventions, our analysis reveals a subtle interconnection between immunity and public health, which together influence the fate of the pandemic. In Section 3 of the Supplementary Material, we have explained why we find the present results to be more realistic. Finally, we have never seen an S-E-I-R framework being used to model a situation such as the complex immune response where a person can be infected exactly twice but not more.

Some of the limitations of the present study are the natural constraints associated with any compartmental or lumped parameter model. For example, the immunity duration used in the model has to be an average over the entire population, which can be refined to some degree by introducing age- or vulnerability-structuring (Sandmann et. al. op. cit). When there are very few cases in a region, the lumped parameter model will no longer be valid. The actual end of the outbreak will be determined by the testing, tracing and treatment of each individual case. Another limitation arises from the current lack of knowledge regarding the human immune response to the new pathogen SARS-CoV-2. Here we have assumed two (plausible) kinds of response, one where immunity completely lapses after a given timeframe and the second where severity-reducing immunity persists indefinitely. Future work may reveal the actual immune response to be more complex than either of these assumptions. Moreover, at least six strains of SARS-CoV-2 have been identified to date (Mercatelli et. al. 2020). At present, we know little about the spreading dynamics of individual strains (Zhang et. al. 2020) and still less about the degree of cross-immunity provided by one strain against another.

In this variability however also lies our model’s primary strength. The model structure makes it easy to incorporate any kind of immune response (Shayak and Mohit 2020). The computational requirement is negligible, with the run for each Notional City taking about one second on a personal computer. Yet, the model is quite powerful since it can generate diverse classes of solutions which are beyond the scope of other models. This means that the moment more information regarding the immune response becomes known, the new information can be encoded into the model framework and accurate case trajectories predicted forthwith. Our analysis will also be of considerable value in calculating the epidemic dynamics after a vaccine is released and mass vaccination is initiated. We shall wait for such an analysis until a vaccine release is imminent or confirmed, since, by that time much more information regarding the immune response to infection and vaccination will be fact instead of speculation.

## Data Availability

There is NO data referred to in the manuscript.

## Acknowledgement

We would like to thank the anonymous reviewer for valuable suggestions which have resulted in substantial improvement to the quality of the presentation.

## Funding statement

We have NO funding to report for this study.

## Conflict of interest statement

We have NO conflict of interest.

## Data Availability Statement

This Article does not use or feature any data.

## SUPPLEMENTARY MATERIAL

### 1 Recapitulation of our prior study

The philosophy behind the delay differential equation model has been proposed in References [S1] and [S2] of this Supplement (cited as References [26,27] in the article proper). We start by summarizing the model derivation and then present the solution classes obtained in these References. This is necessary because the core contribution of the present article adapts this derivation and starts from the same solution classes.

#### Derivation

The transmission of COVID-19 arises as a result of interactions between cases at large in society and healthy people. These interactions may be either direct or via objects. It is described by the following “word equation” :

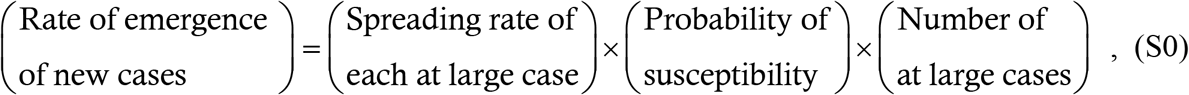

which we intend to represent mathematically.

The first term on the above right hand side (RHS) is itself the product of two quantities : the rate *q*_0_ at which each at large case interacts with other people, and the probability *P*_0_ that an interaction between a case and a susceptible person actually results in a transmission. *q*_0_ is determined by the level of social mobility (lockdown/unlock) and by the degree of physical separation being practised; *P*_0_ depends on whether either or both parties are wearing masks and whether they are washing hands, not touching their face, sanitizing objects etc. Both *q*_0_ and *P*_0_ depend directly on public health interventions, so we can combine them into a single term *m*_0_ which represents this aspect of the disease.

The second term on the RHS of (S0) is the probability that the person with whom the at large case interacts, i.e. a random person in the region, is actually susceptible. For now we consider the case where immunity is permanent. In this case, the probability of a random person’s being insusceptible is the total number of recovered cases divided by the total number of people in the region. The former is approximately *y*, and the latter is *N*. The recovery count is actually a little less than *y* since cases take a finite time to recover. But if the recovery duration is much shorter than the overall epidemic duration, which it is in practice, then we can make the assumption of instantaneous recovery and treat it as *y*. With this assumption, the probability of a random person’s being insusceptible is *y*/*N* and the probability of his/her being susceptible is 1−*y*/*N*.

The structure of the third term arises from the following argument : if all cases take a time *τ* days to recover, and remain at large the whole time, then the number of at large active cases now is exactly the number of people who fell sick between now and *τ* days back – mathematically, this is expressed as *y*(*t*)−*y*(*t*−*τ*). For a maximally realistic picture, we partion the cases into three classes : contact traced cases, untraced symptomatic cases and untraced asymptomatic cases. By definition of the model parameters, the first class amount to fraction 1−*μ*_3_ of the total cases; if the contact tracing starts from freshly reporting symptomatic cases, then the average duration these cases remain at large is *τ*_2_/2 where *τ*_2_ is the transmissible latency period. Untraced symptomatic cases account for fraction *μ*_3_(1−*μ*_1_) of the total cases and they remain at large for the latency period *τ*_2_ before manifesting symptoms and going into quarantine. Finally, untraced asymptomatic cases account for fraction *μ*_3_*μ*_1_ of the total cases and these remain at large for the asymptomatic infection period *τ*_1_. Using the *y*(*t*)−*y*(*t*−*τ*) argument on each class leads to the mathematical form of the third term on the RHS of (S0) as

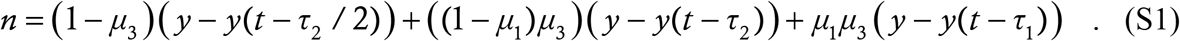

Multiplying all the terms on the RHS of (S0) and simplifying the algebra in (S1), we get

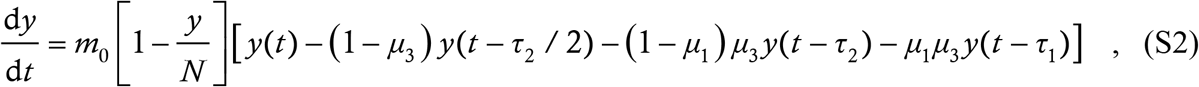

which is the retarded logistic equation proposed in References [S1,S2]. For further details regarding the derivation of (S2), including justification of the approximations involved, we must refer to [S1,S2].

#### Solution classes

Here we present the six solution classes to the model which we have found in [S1,S2], and which form the basis for the solution classes considered in the article proper. For each class, we have run the model in a Notional City of initial susceptible population 3,00,000 as we have done in the article proper. As in there, for each plot we show *y* in blue, 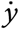 in green and the epidemiological curve as grey bars.

The first solution class, City A, corresponds to elimination of the disease via non-pharmaceutical interventions alone. Reduction of social interactions, masking, contact tracing, testing and similar measures ensure that the reproduction number *R* is less than unity at the start of the outbreak (we have taken it as 0·89 in this particular example) and remains so throughout. Such situations have occurred in New Zealand and Australia.

**Figure S1:**
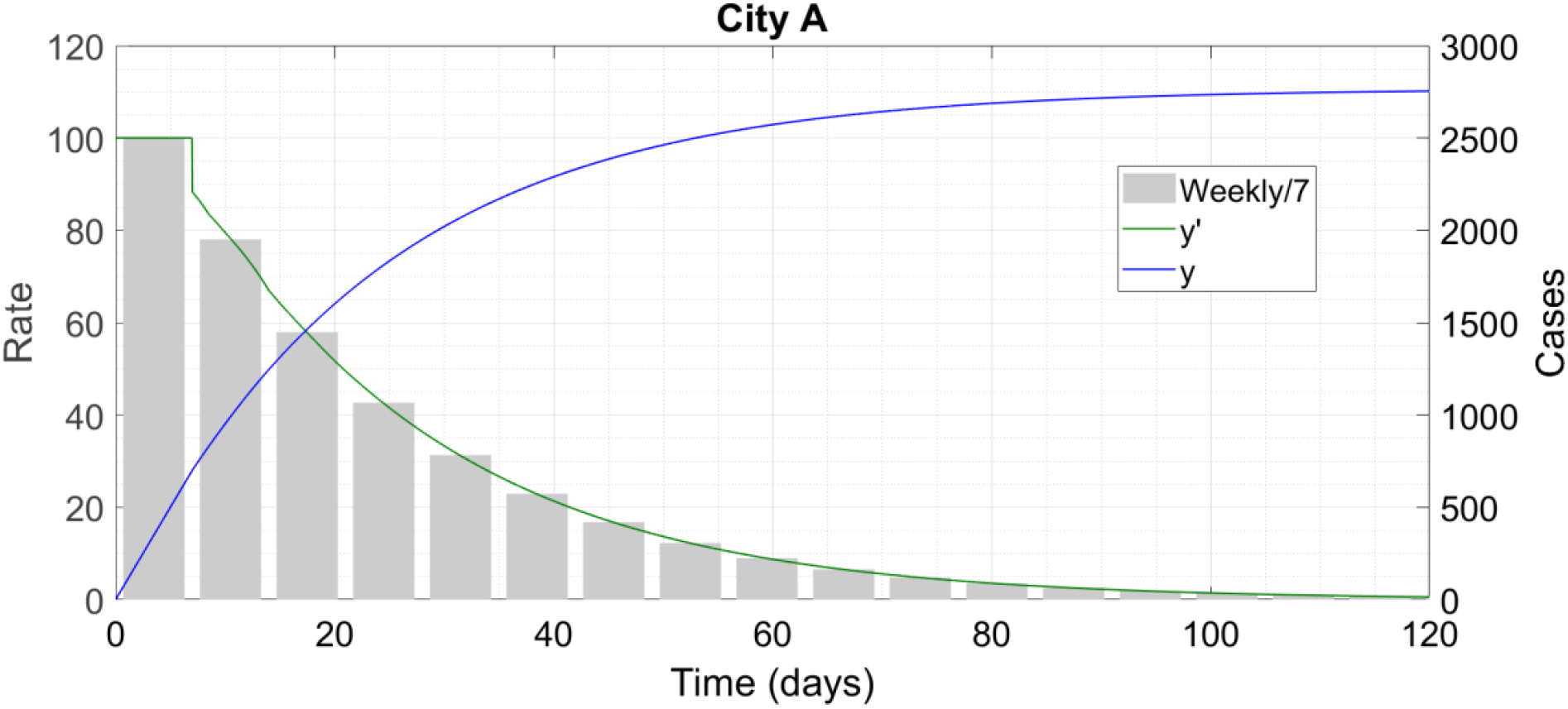
City A contains the epidemic in time.

The second solution class, City B, features non-pharmaceutical interventions at a lower level than City A, which ensures that *R*_0_=1·15. Initially, the outbreak grows exponentially, but then evolves more slowly and infects less than 30 percent of the population by the time it ends. This class of solution (at least in part) has been seen in many regions of the world, such as the cities of Mumbai in India and Buenos Aires in Argentina.

**Figure S2:**
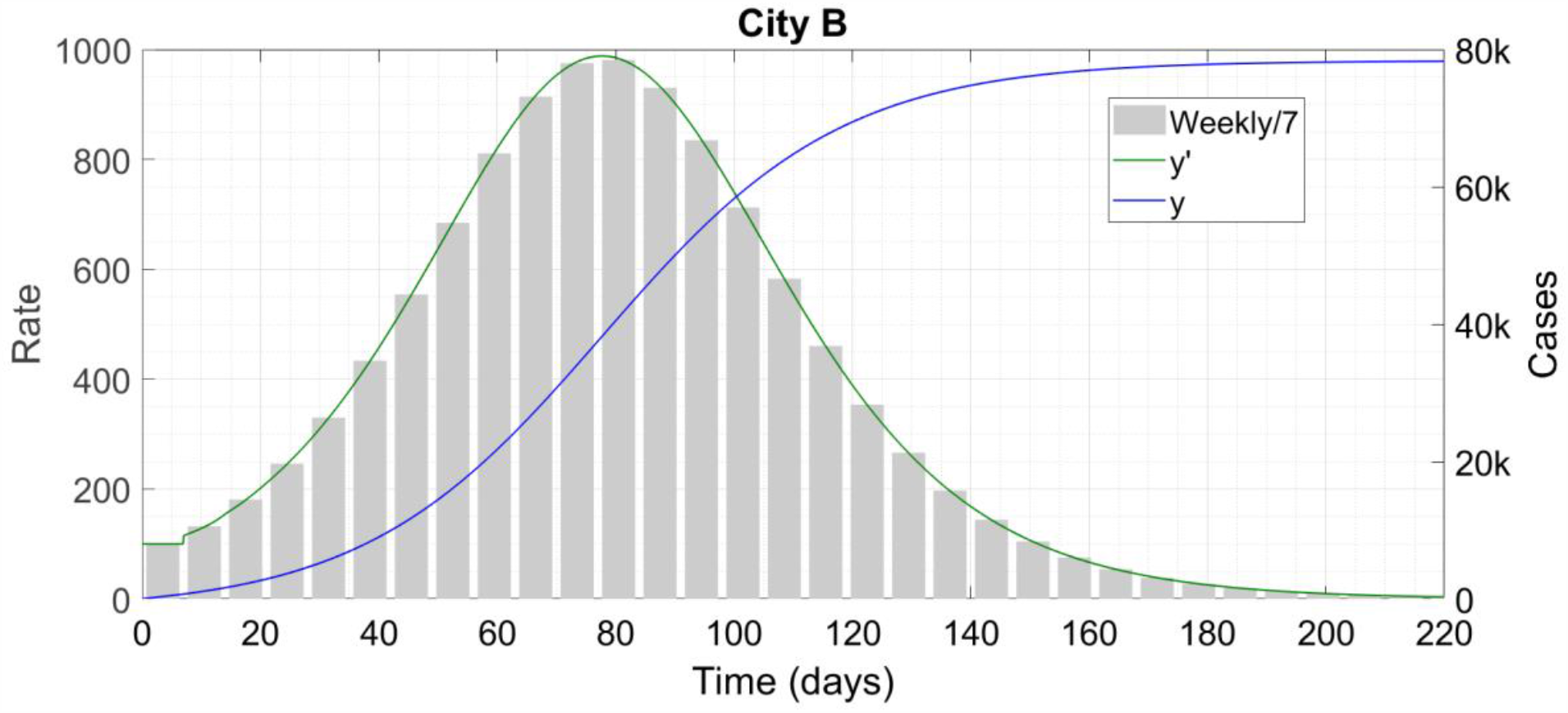
City B has a classical bell-shaped epi-curve.

The third class, City C, attempts no interventions at all. *R* starts off at above 2·5 and remains high throughout until herd immunity is attained. There is very high case count in a very short time. This type of solution has not been seen anywhere in the real world since no local or national authority is so negligent as to adopt a do-nothing course of action.

**Figure S3:**
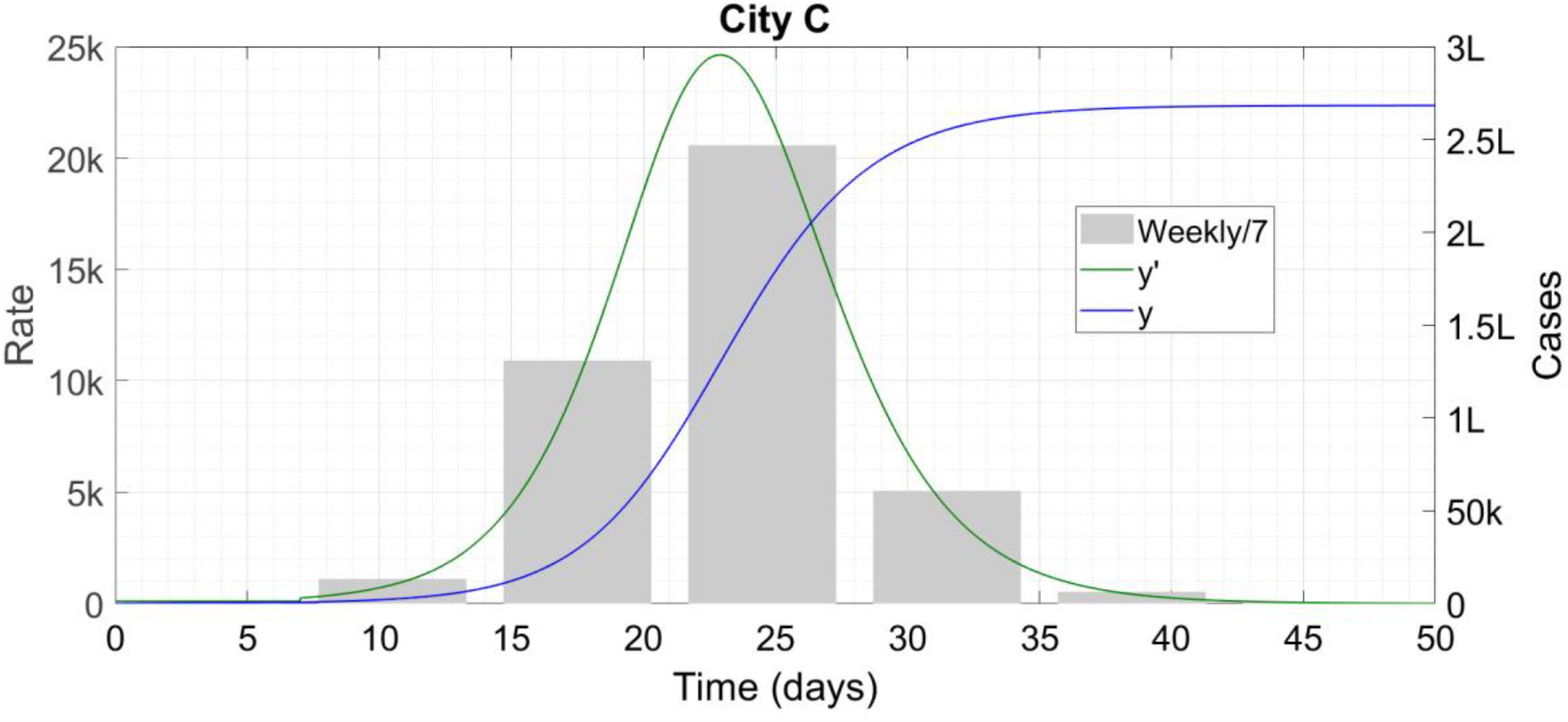
City C proceeds quickly to herd immunity.

The fourth class, City D, shows the effects of a late lockdown. Beginning like City C, this city imposes restrictions at the level of City B when the case count becomes 40,000. The solution combines features of Cities B and C – both the case count and the epidemic duration are intermediate between the two extremes. The first wave of COVID-19 in New York City, USA was of this type.

**Figure S4:**
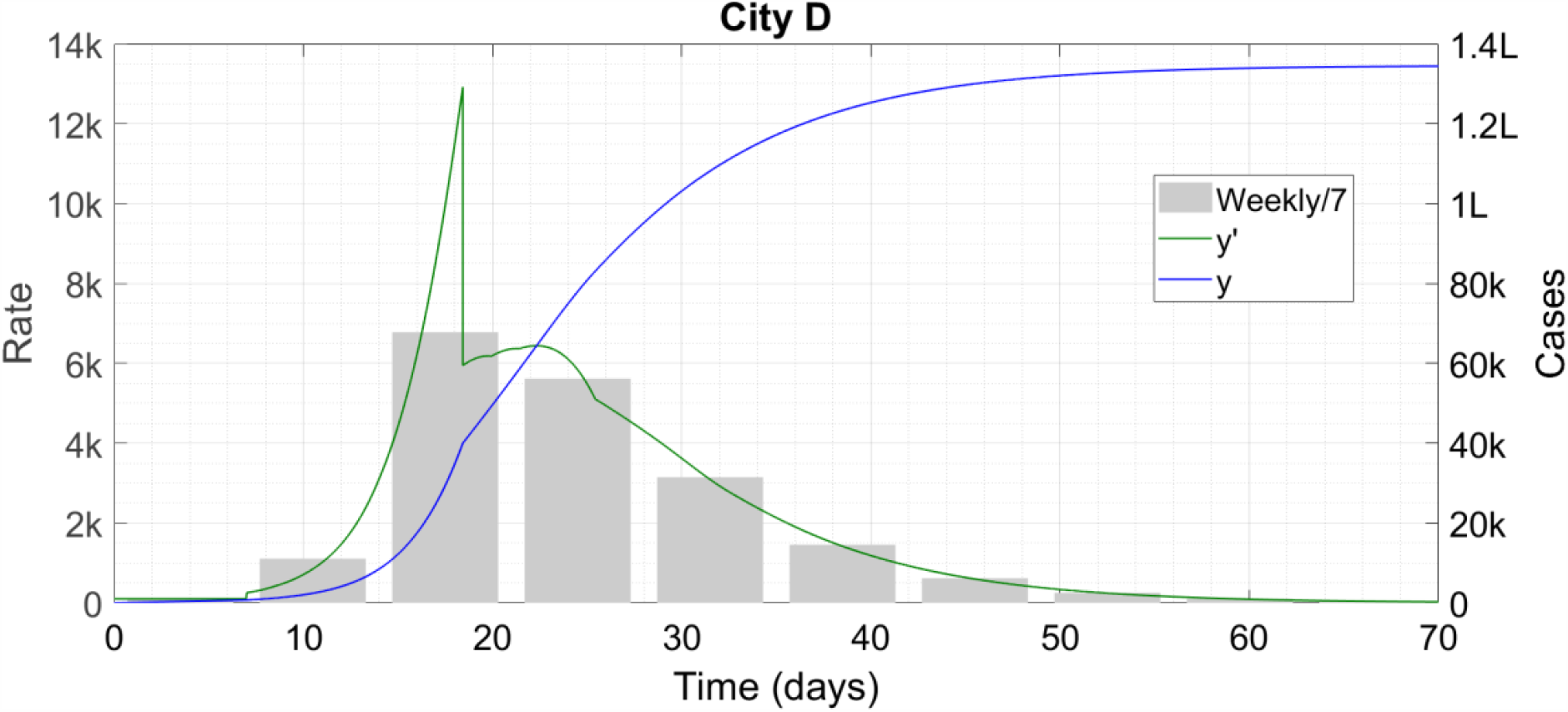
City D combines features of B and C.

Finally, Cities E and F demonstrate reopening. Both start off like City A and then reopen on the 80^th^ day. City E is a success story where increased mobility is combined with high contact tracing, testing etc to keep *R* < 1 throughout. Such reopenings may have taken place in certain East Asian countries.

**Figure 5:**
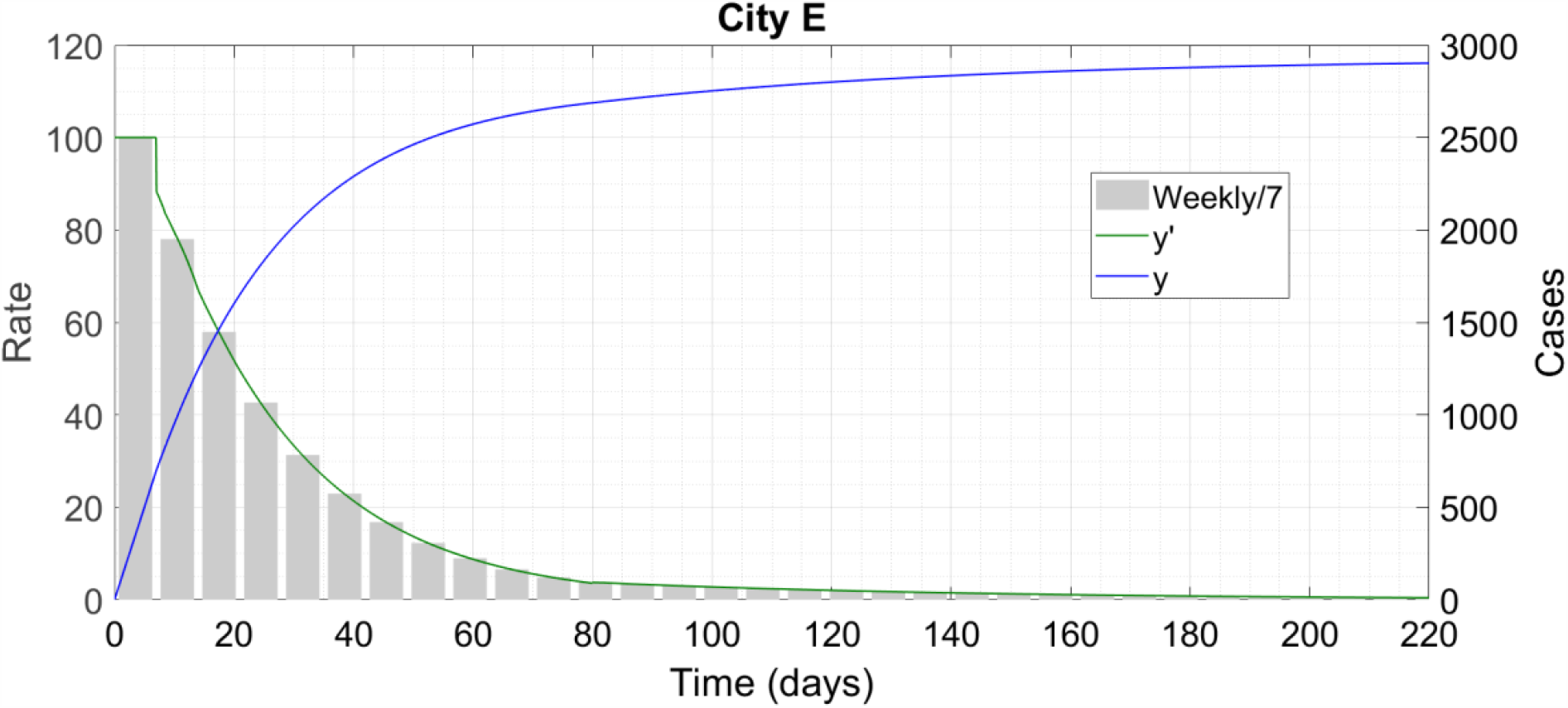
City E is similar to A despite reopening.

City F is a failure story where the increased mobility results in *R* exceeding unity and there is a wave of disease after the 80^th^ day. Reopening driven second waves (with the first wave resembling City B rather than A) have been seen in many parts of the world, especially in USA and Western Europe.

**Figure S6:**
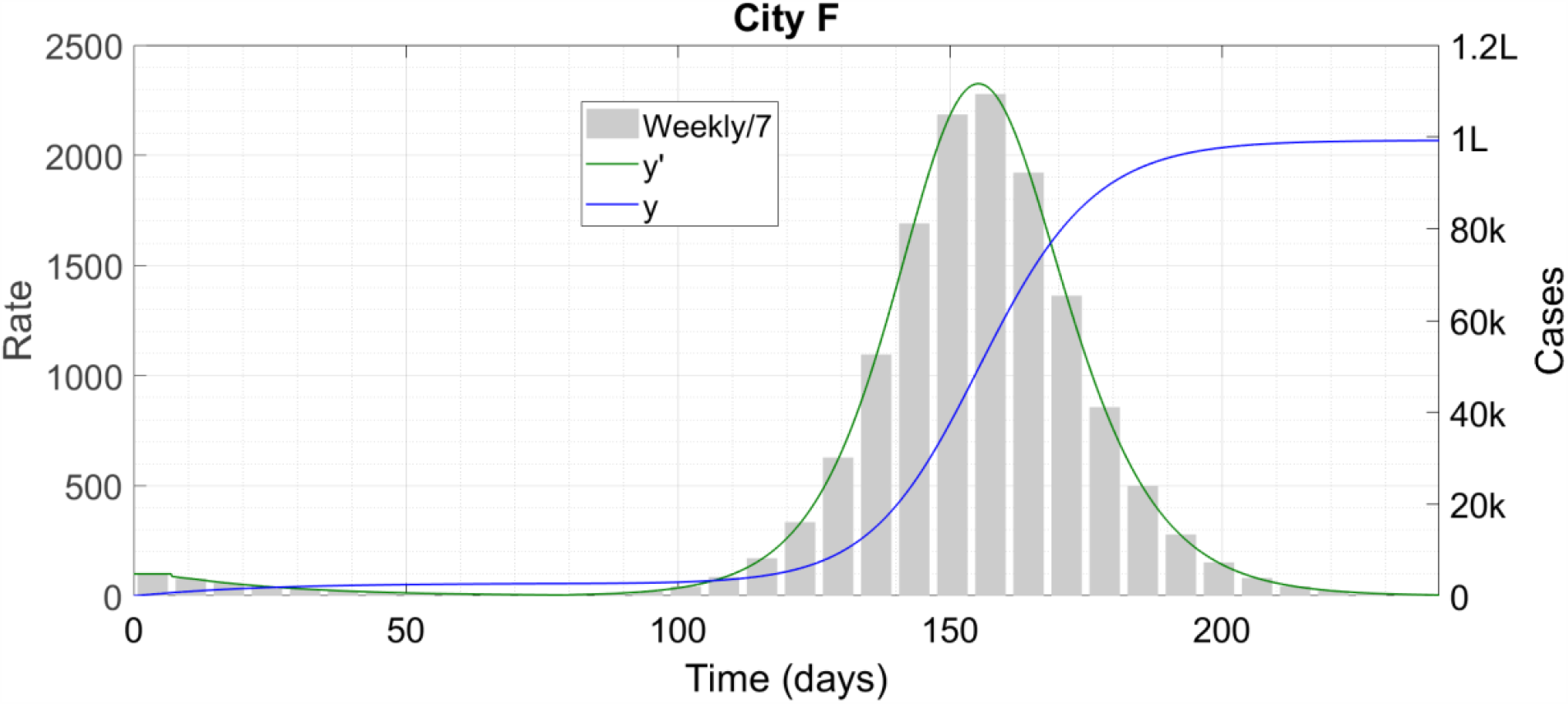
City F shows reopening gone wrong.

This completes the recapitulation of our prior study [S1,S2]. We note that our model, and hence the results of Cities A through G, are more appropriate for an individual city or even a neighbourhood rather than an entire country. This is because the model assumes good interaction among the region’s inhabitants – this assumption is unlikely to hold for a country as a whole, especially under the present circumstances. Rather, a country is made up of dozens of these A-G type cities and the final curve of cumulative cases for this country will be a superposition of the different infection profiles of these cities. Hence, on a national level, public health officials should not attempt to formulate uniform epidemic policies for the country as a whole. It is not uncommon that a phenomenon observed in one region may not be observed somewhere else.

### 2 Development of the model

We now use the modeling philosophy outlined above to derive the governing equations presented in the article proper. We briefly recapitulate the two kinds of immune response from the article proper. Simple immune response : After contracting the infection, a person remains insusceptible for *τ*_0_ days following recovery and then becomes susceptible again. Complex immune response : After contracting the infection, a person remains insusceptible for *τ*_0_ days, then becomes susceptible to the lower virulence form and after a second infection becomes permanently immune.

We first consider the simple immune response. In the structure (S0), immune response is modelled by the second term on the RHS, so that is the only term which will be different from (S2). The immunity threshold of *τ*_0_ means that at any given instant, the people who are immune are all those who have contracted the infection during the last *τ*_0_ days, and no one else. Hence, the number of insusceptibles at time *t* is exactly the number of new infections which have occurred between time *t*−*τ*_0_ and *t*, which is *y*(*t*)−*y*(*t*−*τ*_0_). In proposing this structure, we have retained the approximation of instantaneous recovery and also have ignored deaths. Since the mortality rate of COVID-19 is fortunately quite low, this second assumption is reasonable as well. So the probability that at time *t*, a random person is insusceptible is [*y*−*y*(*t*−*τ*_0_)]/*N*, and the probability that s/he is susceptible is its complement,

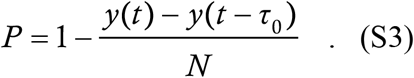

Using this in (S0) immediately gives

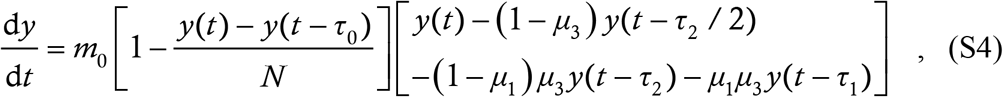

which is (1) of the article proper.

With complex immune response, we need to write the structure (S0) for both *y* and *z*. The public health term *m*_0_ remains the same for both. In the *y*-equation, the second term must denote the probability that a random person is susceptible to the high virulence form. As before, let us count the insusceptibles. Any person who has contracted the high virulence form once is insusceptible to it for all future time, so the number of insusceptibles is *y* and the probability of susceptibility is 1−*y*/*N*. For the third term, we need the total numbers of active and at large *y* as well as *z*, which is (S1) written out for both *y* and *z*. Thus, we get

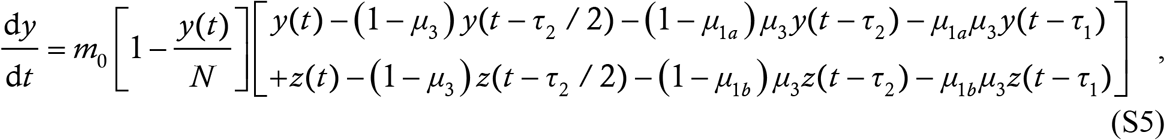

which is (2a) of the article proper.

For the *z* equation, the per-case spreading rate as well as the number of spreaders remain unchanged; the only thing which changes is the probability that a random person is susceptible to the lower virulence form. At time *t*, the only people eligible for contracting the lower virulence disease are those who have already contracted the high virulence form once, and that a time *τ*_0_ or more ago. This is the cumulative number of high virulence infections at time *t*−*τ*_0_, which is *y*(*t*−*τ*_0_). Among these eligible people however, the current count *z*(*t*) have already had the second infection and so must be excluded from the susceptible pool. Thus, the size of the susceptible pool is *y*(*t*−*τ*_0_)−*z*(*t*) and the susceptibility probability is

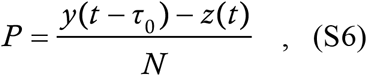

leading to the equation

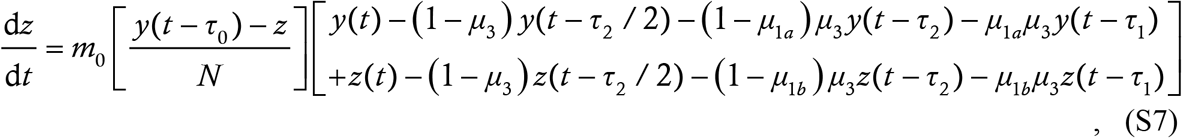

which is (2b) of the article proper. This completes the derivation of the model.

### 3 Qualitative motivation and explanation of the solutions

The different solution classes of (1) and (2) we saw in the article proper are quite counter-intuitive so we motivate them here through some pedagogical examples. We had used one such example in an earlier work [S3] to motivate the City A containment solution, which we now recapitulate briefly. This example was the fantasy kingdom of Coronaland where the king declares a 28-day “100 percent lockdown” – literally nobody is allowed to step out of his/her house. Because Coronaland is fictitious, this does not pose a logistical problem of survival. At the end of the lockdown, every case has either recovered or died at home, and the virus does not exist any longer when normal life is restarted. City A of the article proper is the real-world version of Coronaland since the philosophy is the same – with skilful public health measures, the virus is fed new patients at such a small rate that it itself gets annihilated in time.

The pedagogical examples relevant for temporary immunity are the following. The fictitious city of Coronapur (alla Kanpur, Saharanpur) has an initial population of 500 susceptible people, and has zero cases to start with. The duration of immunity is 200 days and every case recovers within a day (consistent with the modeling assumption above). The spread of the virus here is governed by the following simple rule :

- On any day if there are 5 or more than 5 susceptible individuals present, then exactly 5 of them (randomly selected where necessary) contract the virus
- On any day if there are less than 5 susceptible individuals present then all of them contract the virus

We recall that this is a pedagogical example and not a real-world epidemic spreading dynamic ! The case trajectory of Coronapur is simple enough to predict. There are total 5 cases on day 1, 10 on day 2, 15 on day 3 and so on, all the way upto 500 on day 100. Then, since the first batch of cases is still well within its immunity period, there are no more susceptible people left and the propagation stops. On day 100, the last batch of cases also clears the virus which then does not exist in Coronapur any longer. Everyone has been infected but the virus is dead and the epidemic over.

The fantasy city of Coronabad (alla Hyderabad, Allahabad) has everything same as Coronapur except that the number 5 is replaced by 2. The case trajectory starts off with 2 cases on the first day, 4 on the second, 6 on the third, 200 on the 100^th^ (unlike the 500 of Coronapur) and so on upto 400 cases on the 200^th^ day. Thus, on day 201, there are 100 people who are yet to contract the infection. But, on this day, the cases of day 1 lose their immunity and also get added to the susceptible pool. So the two new infections of day 201 can be anyone from this pool of 102. Similarly on day 202, the two cases from day 201 get removed from the susceptible pool but the two recoveries from day 2 get added and the pool size does not change. This scenario remains invariant for all time – every day there is a susceptible pool of 102 and every day there are 2 new infections from amongst this pool. Even though the initial growth rate is much slower than in Coronapur, the city of Coronabad becomes a land of immortal virus.

The above is exactly what we see in the solutions of (1) and (2). Just as City A is a realistic version of Coronaland, Cities C (also F) and B are realistic versions of Coronapur and Coronabad respectively. A high *R* as in Cities C and F leads to a fast spreading of the epidemic with high initial case counts but puts the progress of the disease to a stop in time. A lower *R* (but of course greater than unity) as in City B leads to slower spreading with lower initial case counts but the disease keeps on perpetuating itself. These examples help us to qualitatively understand the solutions and also convince us that our results are a more accurate representation of reality than literature items (indicated in the article proper) which do not find this complex dependence on *R*.

## Notes

### Competing Interest Statement

The authors have declared no competing interest.

### Funding Statement

We have received NO funding for this study.

### Author Declarations

This research does not involve human or animal subjects so no IRB approvals/exemptions required.

### Summary of Updates

Added a significant literature review at the beginning. Also incorporated suggestions received during peer review, including extension of the supplement.

## References

Abu-Raddad LJ, Chemaitelly H, Ayoub HH, Al Kanaani Z, Al Khal A, Al Kuwari E, et al. Assessment of the risk of SARS-CoV-2 reinfection in an intense re-exposure setting.MedRxiv 2020. available at https://www.medrxiv.org/content/10.1101/2020.08.24.20179457v1.

Ackerly DC. My patient caught Covid-19 twice. So long to herd immunity hopes? Vox 2020. available at : https://www.vox.com/2020/7/12/21321653/getting-covid-19-twice-reinfection-antibody-herd-immunity.

Anna F, Goyard S, Lalanne AI, Nevo F, Gransagne M, Souque P, et al. High seroprevalence but short-lived immune response to SARS-CoV-2 infection in Paris. MedRxiv 2020. available at https://www.medrxiv.org/content/10.1101/2020.10.25.20219030v1.

Bjørnstad O, Shea K, Krzywinski M, Altman N. The SEIRS model for infectious disease dynamics. Nature Methods 2020;17 (6) : 557–8.

CGTN. Ecuador confirms first case of COVID-19 reinfection. Available from: https://news.cgtn.com/news/2020-08-30/Ecuador-confirms-first-case-of-COVID-19-reinfection-Tn8RnpNU40/index.html.

Cooke KL, Van Den Driessche P. Analysis of an SEIRS epidemic model with two delays. Journal of Mathematical Biology 1996; 35 (2) : 240–60.

Crawford KH, Dingens AS, Eguia R, Wolf CR, Wilcox N, Logue JK, et al. Dynamics of neutralizing antibody titers in the months after SARS-CoV-2 infection. MedRxiv 2020. available at https://www.medrxiv.org/content/10.1101/2020.08.06.20169367v1.

Edridge AWD, Kaczorowska JM, Hoste AC, Bakker M, Klein M, Jebbink MF, et al. Human coronavirus reinfection dynamics: lessons for SARS-CoV-2. MedRxiv 2020. available at https://www.medrxiv.org/content/10.1101/2020.05.11.20086439v2.

Giordano G, Blanchini F, Bruno R, Colaneri P, Di Filippo A, Di Matteo A, et al. A SIDARTHE model of COVID-19 epidemic in Italy. arXiv preprint 2020. article number 2003.09861

Goldman J, Wang K, Röltgen K, Nielsen S, Roach J, Naccache S, et al. Reinfection with SARS-CoV-2 and Failure of Humoral Immunity: a case report. Medrxiv 2020. available at https://www.medrxiv.org/content/10.1101/2020.09.22.20192443v1.

Gupta V, Bhoyar RC, Jain A, Srivastava S, Upadhayay R, Imran M, et al. Asymptomatic reinfection in two healthcare workers from India with genetically distinct SARS-CoV-2. Clinical Infectious Diseases 2020. online ahead of print.

Kiran R, Roy M, Abbas S, Taraphder A. Effect of population migration and punctuated lockdown on the spread of SARS-CoV-2. arXiv preprint 2020. article number 2006.15010.

Kosinski RJ. The Influence of time-limited immunity on a COVID-19 epidemic: a simulation study. MedRxiv 2020. available at https://www.medrxiv.org/content/10.1101/2020.06.28.20142141v1.

Kowitdamrong E, Puthanakit T, Jantarabenjakul W, Prompetchara E, Suchartlikitwong P, Putcharoen O, et al. Antibody Responses to SARS-CoV-2 in Coronavirus Diseases 2019 Patients with Different Severity. MedRxiv 2020. available at https://www.medrxiv.org/content/10.1101/2020.09.06.20189480v1.

Lavine JS, Bjornstad ON, Antia R. Immunological characteristics govern the changing severity of COVID-19 during the transition to endemicity. MedRxiv 2020. available at https://www.medrxiv.org/content/10.1101/2020.09.03.20187856v1.

Lumley SF, Wei J, O’Donnell D, Stoesser NE, Matthews PC et. al., “The Duration, dynamics and determinants of SARS-CoV-2 antibody responses in individual healthcare workers,” MedRxiv Article (2020) available at https://www.medrxiv.org/content/10.1101/2020.11.02.20224824v1

McCamon S. USS Roosevelt Sailors Test Positive For COVID-19. Again. NPR Media Article 2020. available at https://www.npr.org/sections/coronavirus-live-updates/2020/05/16/857379338/5-uss-roosevelt-sailors-test-positive-for-covid-19-again.

Mercatelli D, Giorgi FM. Geographic and Genomic Distribution of SARS-CoV-2 Mutations. Frontiers in Microbiology 2020; 11 : 1800.

Mulder M, van der Vegt DSJM, Munnink BBO, van Kessel CHG, van de Bovenkamp J et. al., “Reinfection of SARS-CoV-2 in an immunocompromised patient,” Clinical Infectious Diseases 2020; online ahead of print.

Ogega CO, Skinner NE, Blair PW, Park H-S, Littlefield K, Ganesan A, et al. Durable SARS-CoV-2 B cell immunity after mild or severe disease. MedRxiv 2020. available at https://www.medrxiv.org/content/10.1101/2020.10.28.20220996v1.

Post N, Eddy D, Huntley C, van Schalkwyk MC, Shrotri M, Leeman D, et al. Antibody response to SARS-CoV-2 infection in humans: a systematic review. MedRxiv 2020. available at https://www.medrxiv.org/content/10.1101/2020.08.25.20178806v1.

Robertson LJ, Moore JS, Blighe K, Ng KY, Quinn N, Jennings F, et al. SARS-CoV-2 antibody testing in a UK population: detectable IgG for up to 20 weeks post infection. MedRxiv 2020. available at https://www.medrxiv.org/content/10.1101/2020.09.29.20201509v1.

Sandmann F, Davies N, Vassall A, Edmunds WJ, Jit M. The potential health and economic value of SARS-CoV-2 vaccination alongside physical distancing in the UK: transmission model-based future scenario analysis and economic evaluation. MedRxiv 2020. available at https://www.medrxiv.org/content/10.1101/2020.09.24.20200857v1.

Saplakoglu Y. Recovered patients who tested positive for COVID-19 likely not reinfected. Live Science 2020. available at : https://www.livescience.com/coronavirus-reinfections-were-false-positives.html.

Selhorst P, van Ierssel S, Michaels J, Marien J, Bartholomeeusen K et. al., “Symptomatic SARS-CoV-2 reinfection of a health care worker in a Belgian nosocomial outbreak despite primary neutralizing antibody response,” MedRxiv 2020. available at https://www.medrxiv.org/content/10.1101/2020.11.05.20225052v1.

Sharma MM, Shayak B. Public Health Implications of a delay differential equation model for COVID 19. KIML Workshop Proceedings; KDD 2020. available at https://aiisc.ai/KiML2020/papers/KiML2020_paper_7.pdf.

Shayak B, Sharma MM, Gaur M. A New delay differential equation model for COVID-19. KIML Workshop Proceedings; KDD 2020. available at https://aiisc.ai/KiML2020/papers/KiML2020_paper_6.pdf.

Shayak B and Sharma MM, “A New approach to the dynamic modeling of an infectious disease,” MedRxiv 2020. available at https://www.medrxiv.org/content/10.1101/2020.10.30.20223305v1.

Shrotri M, van Schalkwyk MC, Post N, Eddy D, Huntley C, Leeman D, et al. Cellular immune response to SARS-CoV-2 infection in humans: a systematic review. MedRxiv 2020. available at https://www.medrxiv.org/content/10.1101/2020.08.24.20180679v1.

Siddiqui S, Naushin S, Pradhan S, Misra A, Tyagi A, Looma M, et al. SARS-CoV-2 antibody seroprevalence and stability in a tertiary care hospital-setting. MedRxiv 2020. available at https://www.medrxiv.org/content/10.1101/2020.09.02.20186486v1.

Sinha M. First confirmed Covid reinfection: Have no symptoms, I feel normal, says GIMS nurse. Times of Inida 2020. available at : https://timesofindia.indiatimes.com/city/noida/first-confirmed-covid-reinfection-have-no-symptoms-i-feel-normal-says-gims-nurse/articleshow/78156844.cms.

Tillett RL, Sevinsky JR, Hartley PD, Kerwin H, Crawford N, Gorzalski A, et al. Genomic evidence for reinfection with SARS-CoV-2: a case study. The Lancet Infectious Diseases 2020. online ahead of print.

To KK-W, Hung IF-N, Ip JD, Chu AW-H, Chan W-M, Tam AR, et al. COVID-19 re-infection by a phylogenetically distinct SARS-coronavirus-2 strain confirmed by whole genome sequencing. Clinical Infectious Diseases 2020. online ahead of print.

Trawicki MB. Deterministic Seirs Epidemic Model for Modeling Vital Dynamics, Vaccinations, and Temporary Immunity. MDPI Journal of Mathematics 2017; 5 (1): 7.

Van Elslande J, Vermeersch P, Vandervoort K, Wawina-Bokalanga T, Vanmechelen B, Wollants E, et al. Symptomatic SARS-CoV-2 reinfection by a phylogenetically distinct strain. Clinical Infectious Diseases 2020. online ahead of print.

Wajnberg A, Amanat F, Firpo A, Altman D, Bailey M, Mansour M, et al. SARS-CoV-2 infection induces robust, neutralizing antibody responses that are stable for at least three months. MedRxiv 2020. available at https://www.medrxiv.org/content/10.1101/2020.07.14.20151126v1.

Ward H, Cooke G, Atchison CJ, Whitaker M, Elliott J, Moshe M, et al. Declining prevalence of antibody positivity to SARS-CoV-2: a community study of 365,000 adults. MedRxiv 2020. available at https://www.medrxiv.org/content/10.1101/2020.10.26.20219725v1.

Zhang L, Jackson CB, Mou H, Ojha A, Rangarajan ES, Izard T, et al. The D614G mutation in the SARS-CoV-2 spike protein reduces S1 shedding and increases infectivity. BioRxiv 2020. available at https://www.biorxiv.org/content/10.1101/2020.06.12.148726v1.

Zuo J, Dowell A, Peirce H, Verma K, Long HM et. al., “Robust SARS-CoV-2 specific T-cell immunity is maintained at 6 months following primary infection,” BioRxiv 2020. available at https://www.biorxiv.org/content/10.1101/2020.11.01.362319v1.

## References

[S1] Shayak B, Sharma MM and Gaur M, “A New delay differential equation for the spread of COVID-19,” Proceedings of KiML Workshop, KDD2020, available at https://aiisc.ai/KiML2020/papers/KiML2020_paper_6.pdf

[S2] Sharma MM and Shayak B, “Public health implications of a delay differential equation model for COVID-19,” ibid. (2020) available at https://aiisc.ai/KiML2020/papers/KiML2020_paper_7.pdf

[S3] Shayak B and Rand RH, “Self-burnout – a new path to the end of COVID-19,” MedRxiv Article (2020) available at https://www.medrxiv.org/content/10.1101/2020.04.17.20069443v2

